# Tobacco usage among general Indian population: A meta-analysis of evidence drawn from regional studies between 2010 - 2022

**DOI:** 10.1101/2023.06.11.23291246

**Authors:** Shubhajit Pahari, Diplina Barman, Rounik Talukdar

## Abstract

**Objectives:** The purpose of this review is to generate a national and zonal pooled estimate of Current Tobacco Usage (CTU) in any form, as well as stratified gender and tobacco type (smokeless & smoke) specific estimates among the general Indian population, utilizing evidence from 2010 to 2022.

**Method:** PubMed, Scopus, Science Direct, CINAHL and Google Scholar databases were searched for articles on tobacco use among Indian adults published between January 2010 and October 2022. The NIH Quality Assessment Tool was used to assess study quality, and a random-effects inverse-variance method was used to attain a pooled estimate of usage. Heterogeneity was estimated through I^2^ statistics and prediction intervals, and further subgroup analysis and meta-regression were conducted. To estimate publication bias egger’s test was performed and a leave-one-out analysis was done to establish the sensitivity of our overall pooled estimate.

**Results:** CTU of any form amongst the Indian population irrespective of age group was 35.25% (Confidence Interval (CI) 25.27 - 45.92, I^2^ = 99.7, P-value < 0.001) between the years 2010 - 2022, whereas through 2016 – 2022 regionally drawn estimate was 44.38% (CI 30.57 – 58.64, I^2^ = 99.8, P-value < 0.01). The region-wise highest prevalence was found in the East zone (55.43%) followed by North – East with 51.88% consumption. Though residual heterogeneity was present after subgroup analysis, Variability in estimates was statistically significant by administrative zones and gender-wise consumption (test of subgroup difference P value <0.0001). Leave-one-out analysis proved consistency in our overall CTU estimate.

**Conclusion:** The differences between national-level surveys and regional estimates are highlighted in this review and thereby warrant more regionally representative surveys of tobacco usage for tailor-making prevention efforts alongside, increased regional efforts, improved community-level advocacy, and more coordinated and stringent tobacco prevention policy implementation at national and state levels.

## Introduction

Tobacco dependence is one of the most serious public health risks, with consequences that go beyond cancer, causing a slew of severe debilitating diseases involving the heart, lungs, kidneys, and other organs. Tobacco use is responsible for approximately 33% of cancer-related deaths worldwide, while in India, tobacco-related cancers attributed to 27% of the nation’s cancer burden in 2020.[1, 2] According to the World Health Organization (WHO), there would be more than 8 million tobacco-related deaths per year by 2030, accounting for 10% of all yearly fatalities globally.[3] Tobacco is used by around 32.7% of males and 6.62% of women aged 15 years or more worldwide.[4] Southeast Asian Region (SEAR) recorded the highest Tobacco Usage prevalence in the globe in 2020, which was around 27.9%. The average prevalence among men and women was 46% and 9.7%, respectively.[5] According to the Global Adult Tobacco Survey (GATS 2), 28.6 % (266.8 million) of adults in India aged 15 and above now use tobacco in any form. Studies show that cigarette smoking at a younger age is connected with increased nicotine dependency and long-term tobacco usage.[6] Accounting for other illnesses caused by tobacco use, it is responsible for 14% of all NCD deaths in people aged 30 years and above worldwide,[7] while it is responsible for 9.5% (1 million) of all deaths in India.[8] In 2017-18, tobacco-related direct medical costs accounted for around 5% of total private and public health expenditures in India. The smoking-related expenditures were 74%, whereas the Smokeless tobacco (SLT) related expenses were 26%. Men bore 91% of the overall economic burden, while women bore the remainder.[9]

In response to the significant burden of tobacco use on the country in 2003 Government of India enacted the Cigarette and Other Tobacco Products Act (COTPA), which forbids advertising and governs the trade, commerce, manufacturing, supply, and distribution of cigarettes and other tobacco products in India. With this, the GOI launched the National Tobacco Control Programme (NTCP) in 2007-08 to raise awareness about the negative effects of tobacco, limit tobacco product manufacturing and supply, and guarantee the efficient execution of the COTPA act. After more than 15 years of NTCP implementation, we must assess the current status of tobacco use to inform policy choices concerning the importance of smoking to public health. Though several regional studies and nationalized surveys are available, reports have shown considerable differences between nationwide estimates and varying tobacco usage in different states and union territories.[10, 11]

In this review, we have tried to estimate a nationwide pooled estimate of Current Tobacco Usage (CTU) alongside gender and types of tobacco-specific usage statistics, by drawing data from regional and pan-India studies available between 2010 - 2022. Furthermore, we demonstrated how CTU varied across different administrative zones of India and in the last seven years (2016 – 2022) from GATS 2 survey conduction period till date.

## Methodology

The meta-analysis was registered in the Open Science Framework Database (bearing Registration id: doi.org/10.17605/OSF.IO/P4A9V) after its conceptualization stage. Any deviation from the primary registration has been described. This article adheres to PRISMA & MOOSE guidelines, which is a preferred reporting guideline for meta-analysis of observational studies,[12] we have attached the MOOSE checklist as **Supplementary material 2**.

### Strategies for search

A systematic search was made among five digital databases including PubMed, Scopus, Science Direct, Google scholar and CINAHL on 31/10/2022, to gather evidence relevant to our objective published between January 2011 to October 2022. We did not include studies that have been published beyond 2010 to make our review relevant to today’s changing policy and lifestyle scenario. The rationale behind choosing five digital databases aligns well with the recommended use of a minimum of two databases as charted in AMSTAR guidelines.[13] We crafted our search strategy utilizing a combination of MeSH headings, major topics and indexed keywords. The central theme of our search strategy has been described below:

((“prevalence”[Title/Abstract]) OR (“epidemiology”[Title/Abstract]) OR (“use*”[Title/Abstract]) OR (“frequency”[Title/Abstract]) OR (“occurrence”[Title/Abstract]) OR (“incidence”[Title/Abstract]) OR (“survey”[Title/Abstract])) AND ((“tobacco”[MeSH Major Topic]) OR (“smoking”[MeSH Major Topic]) OR (“cigarette smoking”[Title/Abstract]) OR (“tobacco”[Title/Abstract]) OR (“cigar smoking”[Title/Abstract]) OR (“smok*”[Title/Abstract]) OR (“smokeless tobacco”[Title/Abstract]) OR (“smoking behavior”[Title/Abstract]) OR (“smoking habit”[Title/Abstract]) OR (“beedi”[Title/Abstract]) OR (“hooka”[Title/Abstract]) OR (“chhutta”[Title/Abstract]) OR (“dhumti”[Title/Abstract])) AND ((“india”[MeSH Major Topic]) OR (“india”[Title/Abstract]))

The search strings were manually translated pertaining to the need of other individual databases. For google scholar, we kept our search most relevant to our objective by using the “sort by relevance” option in the search engine, and first ten web pages were considered for scrutiny. Further we manually searched the reference lists of identified articles which were eligible for full text screening for probable inclusion in our review.

### Selection Criteria

Our selection criteria were based on the **CoCoPop** approach, which has been advocated as a preferable method in systematic review and metanalysis of prevalence/ proportions.[14] The framework was described as **Co**ndition (CTU* of any type – smoke or smokeless form), **Co**ntext (India, evidence drawn from published peer-reviewed studies through 2010 – 2022), and **Pop**ulation (Adult general Indian population). Studies adhering to the below inclusion criteria were considered: 1. Observational/ Community-based prospective and cross-sectional studies enrolling participants from the 15+ age group. We considered the 15+ age group as adults similar to most of the nationalized surveys.[15] 2. Studies with adequate data in the public domain for extraction of tobacco usage proportion. 3. As this review concentrate on India, published articles only in the English language were included which holds the status of the associate official language of India. Articles originating from India in medicine and related field are chiefly in English language.

*CTU is defined in this review as – current user (used tobacco in the preceding 1 to 6 months. / Explored through cotinine test etc.) or explored CTU as current daily usage as per individual study definition.

Exclusion Criteria were: 1. Studies including secondary data (i.e., studies utilizing GATS survey data, NFHS data etc.) the rationale behind this was that we tried to highlight the dissimilarities between Nationalized surveys and regionally representative studies, so we omitted studies that have used nationalized survey data for analysis. 2. Unpublished studies and grey literature because of anticipated insufficiency in reporting data. 3. Studies conducted on any hospital-based or institutionalized population (Outpatient based, Prisons, mental health facilities, etc.) 4. Studies conducted solely on any specific subset of the population or only enrolled participants with any specific attribute (e.g., studies enrolling only one gender, studies on industrial/ construction workers, slum populations, etc.) to avoid unduly skew in usage statistics.

### Study selection

Title screening was done to identify the studies matching the inclusion criteria by two reviewers (DB & SP) independently. Further included studies were imported to Mendeley Reference Manager version (2.77.0) for identifying duplicates. Disagreements if any between the reviewers were resolved through discussion by involving another reviewer RT. Abstract screening of the identified studies was done. The full text was retrieved for selected abstracts and was subsequently reviewed for eligibility and sufficiency of data. A total of thirteen studies were included. The exclusion of the studies was done based on methodological mismatch and predefined criteria and definitions.

### Data extraction

The data extraction form was created in Microsoft® Excel® 2019 MSO (Version 2209 Build 16.0.15629.20152), enlisting information on 1. First author details 2. Publication year 3. Study type & setting 4. Administrative zones, 5. The age range of the population involved 6. sample size 7. CTU 8. Types of tobacco usage (smokeless vs. smoke form) 9. Gender-wise tobacco usage statistics as available in some studies.

Data extraction was conducted by (DB& SP) and was subsequently reviewed by RT.

### Quality assessment

All the finally selected articles were assessed using the NIH quality assessment tool. Each question relevant to included study designs with a satisfactory answer was awarded a score of one, otherwise zero. Questions that were not relevant to the studies were not included and were not counted in the denominator to calculate an individual score. Two reviewers SP & DB in consensus with RT resolved any disagreement about quality assessment. The complete NIH tool can be found on the nhlbi.nih.gov website.[16]

### Data synthesis and analysis

The main outcome of the review was the prevalence of CTU among the general Indian population irrespective of age group, which was described as a proportion with lower and upper confidence limits.

R-studio (version 4.1.3, 2022; The R Foundation for Statistical Computing, Vienna, Austria)[17] was used for conducting the Meta-analysis. A dedicated command package for meta-analysis of single proportions: “Metaprop (R documentation.org was used)” Extracted data was utilized to calculate the standard error (SE) of proportions of tobacco usage. The formula used is depicted below.[18, 19]

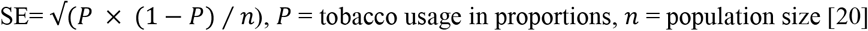

For pooling proportions, a random effect inverse variance method was used which involves the calculation of weighted average using SE, to be supplied to the data frame for the meta prop package in the R-studio. The justification for using a random effect model relates to the fact that a significant amount of study-level heterogeneity in terms of the research population, location, and methodology was anticipated during the study selection and finalization. Heterogeneity was assessed through I^2^ statistics (<25%-Mild, 25-75%-Moderate, >75%-High heterogeneity)[21] and we calculated the prediction interval, which is the range within which we can forecast with 95% certainty that the true prevalence of tobacco usage estimate in a future study will fall.[22] Subgroup analysis was performed based on the year of the study and administrative zones, and gender. The variable year of the study was divided into two groups based on GATS 2 survey conduction year. GATS 2 in 2016 – 2017 assessed tobacco usage amongst the Indian population. So, in our review, we have grouped selected studies in Pre-GATS 2 (2010 – 2015) & 2016 – 2022 periods. Univariate meta-regression was conducted to explore the reason for heterogeneity on the same aforementioned variables. Publication bias was assessed using a Funnel plot by plotting arcsine transformed proportion on the x-axis against sample sizes on the y-axis. Evidence shows that for proportional/ prevalence meta-analysis plotting transformed proportions vs sample size is a better method to assess publication bias.[23] Further asymmetry of the plot was also tested using Egger’s test where a p-value <0.05 was considered statistically significant.[24] We also evaluated the sensitivity of our overall CTU estimate using leave-one-out analysis, which recalculates the effect estimate by eliminating one study at a time and collating the results in a forest plot. This allows us to see the variation in pooled estimates and, as a result, the stability.[25]

## Results

Out of 3803 studies identified through digital databases search, 13 studies matched the inclusion criteria. A complete flowchart adhering to the PRISMA/ MOOSE statement for stepwise selection and inclusion of studies has been presented in **Figure 1**.

**Figure 1:**
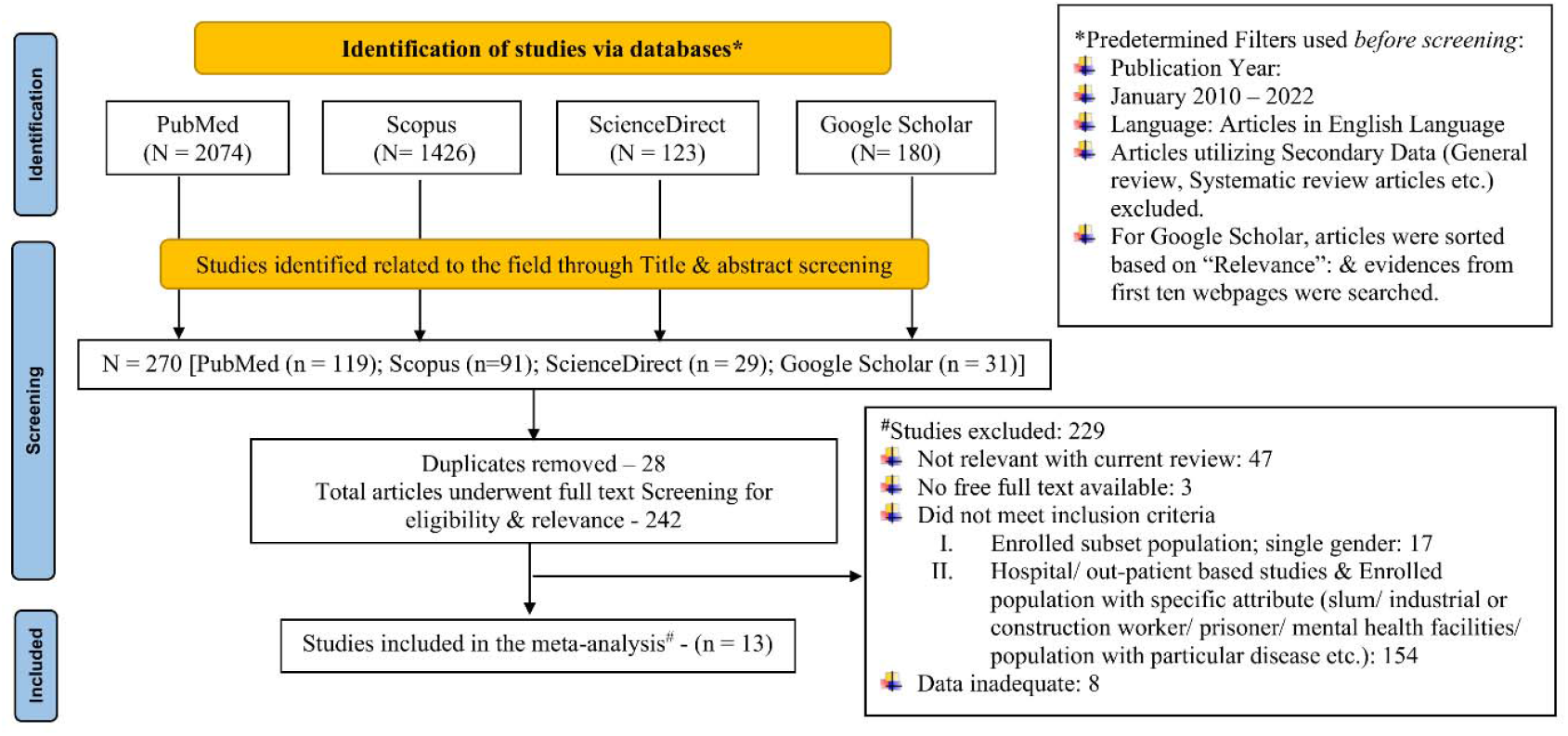
PRISMA flowchart for selection and inclusion of studies in the review

### Characteristics of the studies included

The thirteen studies included a total of 64591 participants from urban, rural, and mixed settings. The median sample size of included studies was 1635, with IQR being 887 (Q_1_) – 6864 (Q_3_). Eight of the included 13 studies were carried out between 2010 and 2015, while the remaining five were conducted between 2016 and 2022. There were three studies each from the country’s north and south zones, as well as two studies from each of the northeast, central, and east zones. We could find only one study based on our inclusion criteria from the west zone. Gender-specific stratified data were available in nine of the thirteen studies included.

**Table 1** depicts the complete study characteristics including study design, settings, population, age groups, Overall & type specific CTU estimate, etc.

**Table 1:** Characteristics of the studies included in the review

| Sl no | Author name & Publication Year, References | Study Conduction Year | Study Type | Study region | study area | Study Setting | Population | Age range | sample size | Overall, Tobacco Users | Tobacco user |  | Types Tobacco |  |  |
| --- | --- | --- | --- | --- | --- | --- | --- | --- | --- | --- | --- | --- | --- | --- | --- |
|  |  |  |  |  |  |  |  |  |  |  | Male | Female | SLT | Smoke | Both |
| 1 | Oswal et al. 2021[40] | 2021 | Cross-sectional | North east | Assam, Nagaland and Meghalaya | Village/ Rural | General Population | <25 - >65 | 1400 | 603 | 447 | 156 | 375 | 211 | 17 |
| 2 | Chockalingam et al. 2013[41] | 2011 | Cross-sectional | South | Chennai | Urban, Semi-urban and rural | General Population | 15 - ≥ 65 | 7510 | 1604 | 1407 | 197 | 589 | 1015 | – |
| 3 | Rautela et al. 2016 | 2016 | Cross-sectional | Central | Shrikot, Uttarakhand | Rural | General Population | 20 - > 60 | 155 | 20 | -- | NA | 7 | 13 | -- |
| 4 | Srivastava et al. 2022[42] | 2018 | Cross-sectional | Central | Uttar Pradesh | Urban and rural | General Population | > 15 | 1461 | 736 | 566 | 170 | 311 | 543 | -- |
| 5 | Giri et al. 2021[43] | 2019 | Cross-sectional | Nort east | Assam | Urban & Rural | General Population | 15 - ≥ 65 | 5000 | 3028 | 1835 | 1193 | 3028 | NA | -- |
| 6 | Hasan et al. 2020[44] | 2017 | Cross-sectional | North | Uttar Pradesh | Rural | General Population | ≥18 - > 60 | 6218 | 3884 | 3753 | 131 | 2669 | 1913 | -- |
| 7 | Agrawal et. al. 2013[45] | 2012 | Cross-sectional study | North | Gorakhpur district, Uttar Pradesh | Urban & Rural | General Population | 15-60 | 1635 | 394 | -- | -- | 299 | 267 | -- |
| 8 | Koothati et al 2017[46] | 2012 | Cross-sectional study | South | Telangana | Urban & Rural | General Population | 16-75 | 3200 | 750 | 554 | 196 | 334 | 320 | 96 |
|  |  |  |  |  |  |  |  |  |  |  | Male | Female | SLT | Smoke | Both |
| 9 | Kumari et al 2020[47] | 2014 | Cross-sectional study | North | Rohtak, Haryana | Urban | General Population | 15-64 | 1440 | 328 | 321 | 7 | -- | -- | -- |
| 10 | Sarma et al 2019[48] | 2016-17 | Cross-sectional study | South | kerela | Urban & Rural | General Population | 18-69 | 12012 | 2438 | -- | -- | -- | -- | -- |
| 11 | Kahar et al. 2016[49] | 2010 - 2015 | Cross-sectional study | West | Gujarat | Rural | General Population | 18 - ≥ 55 | 23953 | 4365 | 3988 | 377 | 2257 | 3398 | NA |
| 12 | Giri et al.2019[50] | 2017 | Cross-sectional study | East | Odisha | Rural | General Population | > 40 | 233 | 150 | -- | -- | -- | -- | -- |
| 13 | Bhattacharya et al. 2020[51] | 2018 | Cross-sectional study | East | Kolkata, West Bengal | Urban/ Rural | General Population | ≥ 15 | 374 | 174 | 140 | 34 | 76 | 98 | |

### Quality assessment

The mean NIH quality assessment score for the included studies (n=13) was 81.70 with a standard deviation of 16.62. Eleven studies scored more than or equal to 75% and were graded as methodologically good following the scoring classification used by Sommer et al in their study.[26] One study was deemed “fair,” while another was deemed “poor,” with a minimum score of 37.5. **Figure 2** depicts the complete quality scoring for each study.

**Figure 2:**
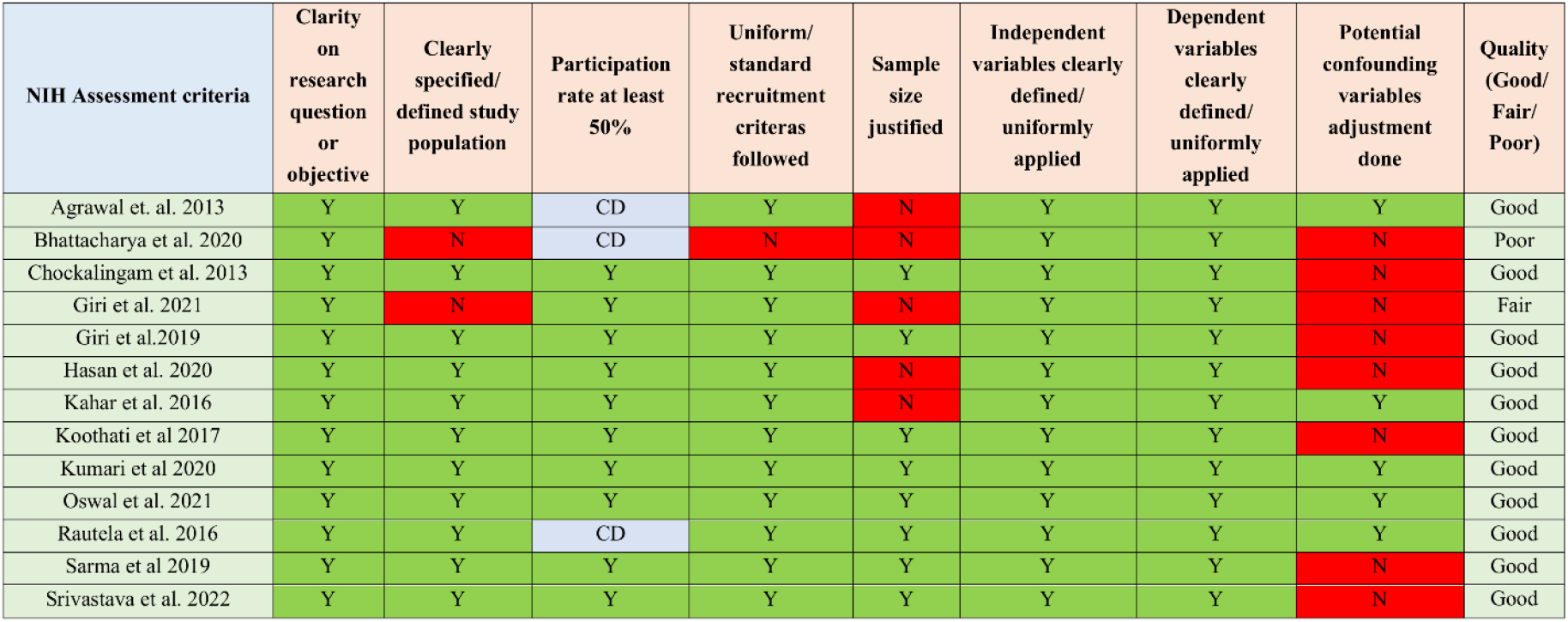
Quality assessment of included studies through NIH Quality assessment tool (Y = Yes/ satisfactory, N = No/ absence, CD = Cannot determine)

### Prevalence of tobacco usage

The overall pooled estimate of any type of CTU among the Indian population of any age group was 35.25% (CI 25.27 - 45.92, I^2^ = 99.7, P-value < 0.001) with a prediction interval between 3.19 – 78.74. **(Figure 3)**

**Figure 3:**
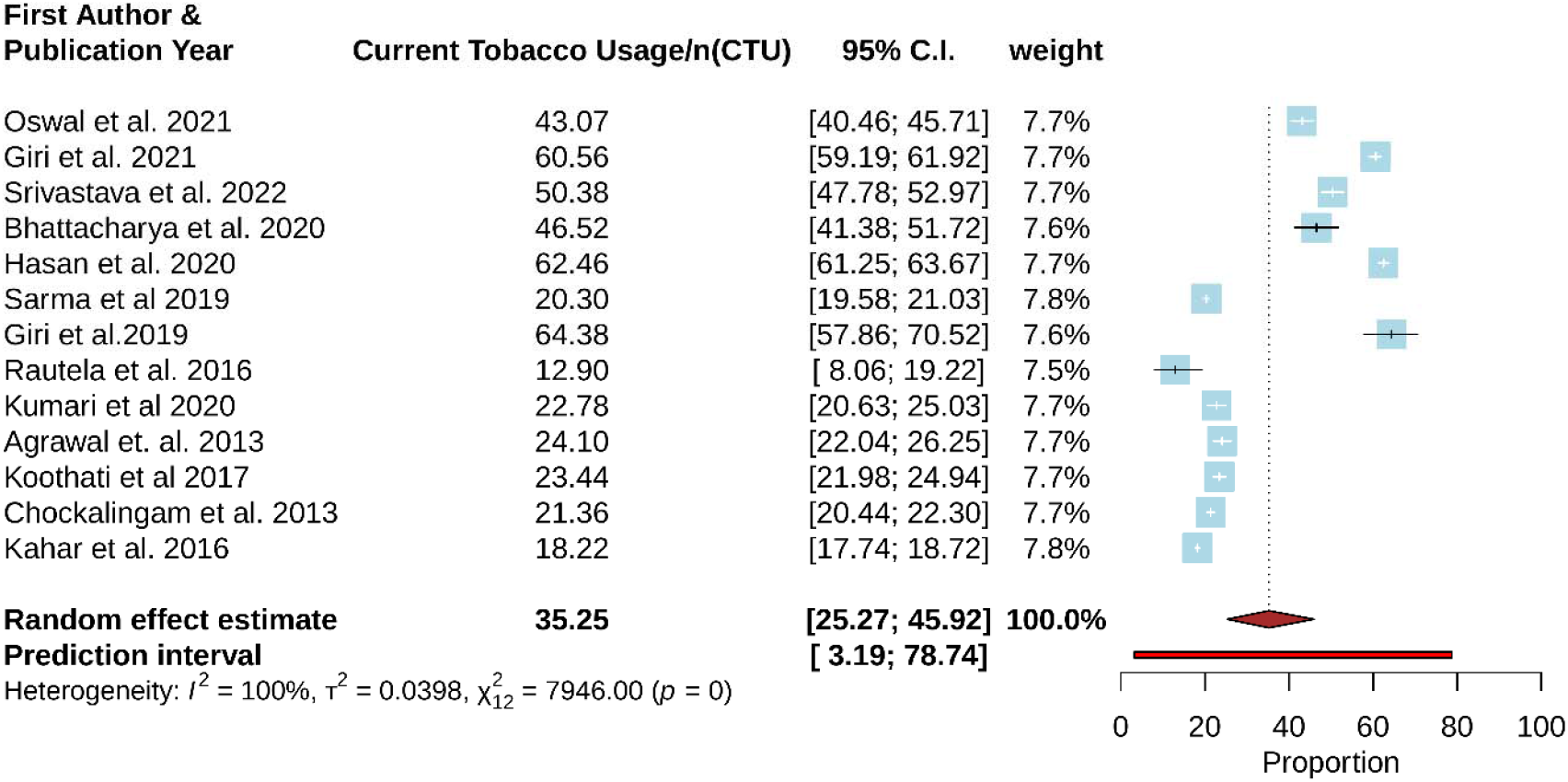
Forest Plot- overall current tobacco usage among general Indian population between period 2010 – 2022

Subgroup analysis in terms of CTU in different administrative zones in India revealed the highest usage in the Eastern part of the country with an estimated 55.43% (CI 37.86 – 72.32, I^2^ = 95%, p-vale <0.01), followed by 51.88% (CI 34.88 – 68.66, I^2^ = 99%, p-vale <0.01) in North-Eastern zone. The lowest CTU was noted in the Southern zone of the country with an estimate of 21.36% (CI 19.88 – 23.36, I^2^ = 87%%, p-vale <0.01). A CTU of 30.03 and 35.75% were estimated in the central and northern zones respectively. **(Supplementary material 1; figure 1)**

Stratified analysis based on gender and type of CTU was done. The overall prevalence pooled from nine studies for any form of CTU noted amongst males was 54.13% (CI 43.56 - 64.51, I^2^ = 99.7, P-value < 0.01), whereas in females the estimated prevalence was 15.08% (CI 6.37 - 26.60, I^2^ = 99.8%, p-value < 0.01). **(Supplementary material 1; Figure 2)**

Data on the use of smoke and smokeless tobacco was provided in nine and ten out of thirteen studies, respectively. The pooled usage of smokeless tobacco from 10 articles was estimated to be 20.89% (CI 11.59 - 32.07, I2 = 99.8%, p-value < 0.001), whereas data from nine studies revealed a tobacco smoking prevalence of 18.91% (CI 13.30 - 25.24, I2 = 99.7%, p-value < 0.01). **(Figure 4(A), & (B))**.

**Figure 4:**
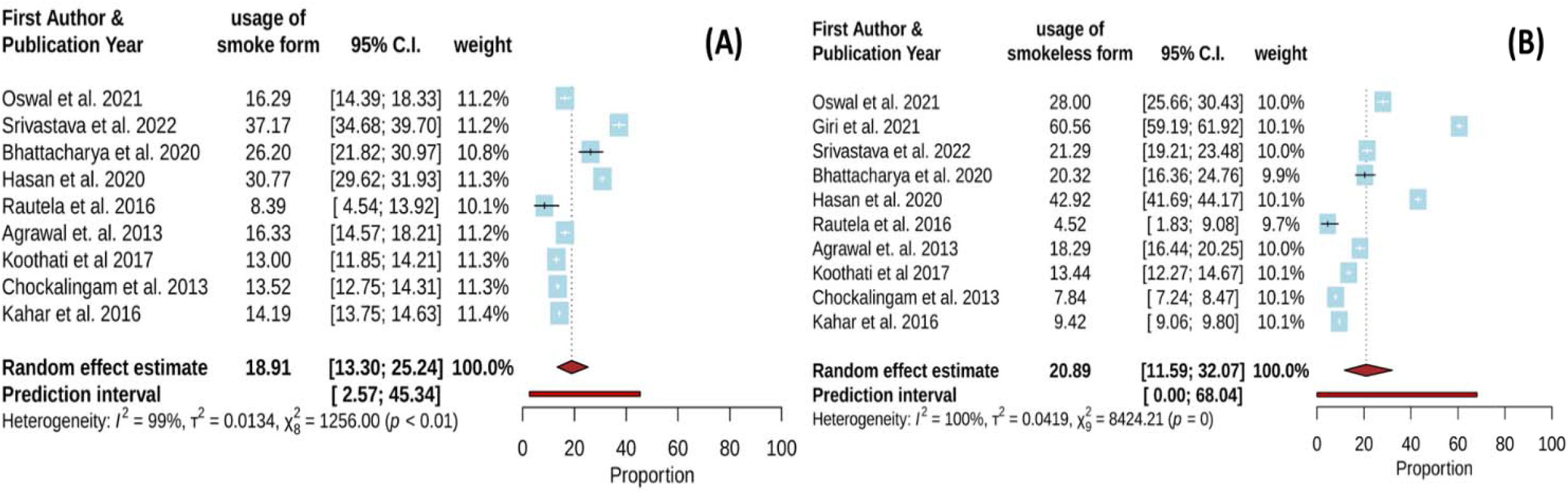
Forest Plot- overall current usage of (A) Smoke form, (B) smokeless form among general Indian population in the period of 2010 – 2022

Further, pooled CTU estimates from eight studies conducted between 2016 and 2022 were 44.38% (CI 30.57 - 58.64, I2 = 99.8%, p-value = 0.001), whereas estimates from five studies conducted between 2010 and 2015 were 21.82% (CI 19.70 - 24.02, I2 = 96%, p-value = 0.001). **(Supplementary material 1; Figure 3)**

According to the test of subgroup differences, there were statistically significant subgroup effects for variables: administrative zone, study year, and gender, with p-values for subgroup differences, reported as < 0.0001, 0.001, and < 0.0001 respectively, **Table 2**). Further Univariate Meta-regression analysis revealed a significant p-value for the moderator variable: year of study and gender (0.009, < 0.0001 respectively). However, there was still unexplained heterogeneity within each subgroup after subgroup analysis. (**Table 2**) As the uneven distribution of studies and inherent heterogeneity majorly contributed to this review, therefore this analysis may not be able to identify differences among subgroups.

**Table 2:** Subgroup analysis based on Gender, administrative zones, and Study conduction year

|  |  | No. Of Studies (n) | CTU (prevalence) | 95% CI | Meta-regression | Heterogeneity |  |  |
| --- | --- | --- | --- | --- | --- | --- | --- | --- |
|  |  |  |  |  | P- Value | I<sup>2</sup> (%) | P- Value | p-value of Test of Subgroup difference |
| Based on Gender (N= 9) | Female | 9 | 15.08 | 6.37 – 26.66 | < 0.0001 | 99.8 | < 0.01 | < 0.0001 |
|  | Male | 9 | 54.13 | 43.56 – 64.51 |  | 99.7 | < 0.01 |  |
| Geographic zones (N=13) | East | 2 | 55.43 | 37.86 - 72.32 | 0.17 | 94.7 | < 0.01 | < 0.0001 |
|  | South | 3 | 21.60 | 19.88 – 23.36 |  | 86.8 |  |  |
|  | North | 3 | 35.75 | 13.43 – 61.99 |  | 99.6 |  |  |
|  | West | 1 | 18.22 | 17.74 – 18.71 |  | -- | -- |  |
|  | North-east | 2 | 51.88 | 34.88 - 68.66 |  | 99.3 | < 0.01 |  |
|  | Central | 2 | 30.03 | 2.75 – 70.20 |  | 99.0 |  |  |
| Based on Study Conduction Year (N=13) | 2010 - 2015 | 5 | 21.82 | 19.70 – 24.20 | 0.01 | 96.0 | <0.01 | < 0.01 |
|  | 2016 - 2022 | 8 | 44.38 | 30.57 – 58.64 |  | 99.8 | < 0.001 |  |

### Sensitivity Analysis

We performed a leave-one-out analysis to confirm the stability of the overall CTU estimate, which inferred that no single study had a substantial influence on the findings, with the pooled CTU estimate ranging between 32.96 and 37.32 (95% CIs between 23.39 and 48.06). The I^2^ index constantly ranged between 99.8% and 99.9%. **(Figure 5(A))**

**Figure 5:**
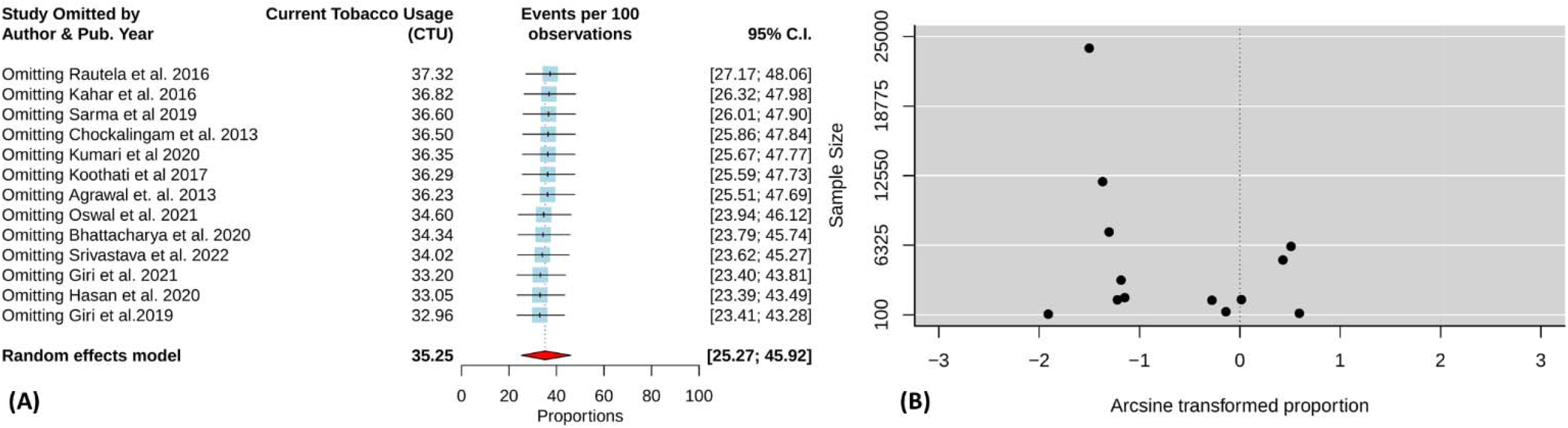
(A) Leave-one-out analysis establishing sensitivity of overall CTU estimate; (B) Assessment of publication bias-funnel plot

### Publication bias

Subjective evaluation of the funnel plot revealed mild asymmetry; however, Egger’s regression test was conducted which elicited a P value of 0.80, indicating the absence of publication bias. **(Figure 5(B))**

## Discussion

Our analysis showed that the overall pooled estimate of CTU among Indians from 2010 to 2022 was 35.25%, with the East (55.43%) and North-East (51.88%) zones having the highest prevalence.

According to the WHO-Global Health Observatory report in 2010 CTU in South Asian countries was 37.3% whereas it declined to 27% in 2020. Our nationalized estimate on current overall usage (35.25%) drawn from literature through 2010 – 2022, fares well against the WHO data, which estimates an overall current usage of 27.2% in India in 2020.[27] Subgroup analysis in the current review shows that before the GATS 2 survey in the period of 2010 to 2015 pooled estimate of CTU in India was 21.82%, whereas in the 2016 - 2022 period it had a considerable increase to 44.38%. However, as per National Family Health Survey 5 (NFHS-5, 2019 - 2020) data, usage of any form of tobacco was estimated as 27.3% among Indian adults,[28] similarly GATS 2 survey (2016 -2017) indicates an overall usage of 28.6% in India.[29] One of the probable reasons for the extreme variation between our 2016 - 2022 CTU prevalence and nationalized survey estimates is the non-uniform representation of studies from different administrative zones in the same period, moreover selected studies in the 2016 – 22 period were mostly from two high CTU prevalence zones i.e. North-east & East.

The other reason behind our increased pooled estimate in 2016 - 22 period (44.38%) in comparison with recent national-level surveys (GATS 2 estimate – 28.6% [29]) is aligned with the fact that multiple studies have cited the stark difference between the reports from national surveys versus regional level estimates.[10, 30, 31] Moreover, if we look at the current NFHS-5 state-level data, there are estimates of tobacco usage amongst men as high as around 72% in Mizoram with a median usage of 39.8 (Range 56) amongst all the states and union territories.[30, 32, 33] Though more studies from different zones are needed to attain a more robust nationalized estimate, our meta-analysis draws data from small-scale regional studies across all of India thereby taking into account the region-specific variations.

Our estimation of any kind of CTU amongst Indian men was 54.13% between 2010 to 2022, which is placed at a higher level than the reported 46% average prevalence of tobacco use among males in the South East Asian Region in 2020.[5] Similarly, GATS-2 survey data revealed usage of 42.4% amongst Indian males.[29] A study published by Sreeramareddy et al. in 2014 estimated the prevalence of tobacco usage in any form amongst Indian men to be 36.8%.[34] They borrowed cross-sectional data from nationally representative surveys which again justifies our estimate taking into account being more regionally representative and being drawn from 12-year-long evidence. More recent reports gathering data from NFHS 2^nd^,3^rd^, and 4^th^ rounds revealed that in 2015 – 2016 prevalence of tobacco usage amongst Indian males was 27.3%, which dropped from 36.81% as estimated during the 2005 – 2016 period.[31]

As per reports, approximately 22% and 9% of women in developed and developing countries, respectively, are daily smokers.[35] In terms of tobacco usage among women in India, GATS-2 and NFHS-5 survey data report 9% and 14.2%, respectively. In this review, CTU among Indian women was estimated to be moderately high at 15.08%. A study on national survey data by Goel et al. found a 5-year doubling rate of women smoking prevalence in India. (In 2010 2.9%, up from 1.4% in 2005).[36] An increase has also been noted in recent NFHS data concerning GATS 2 data. Even in rural India tobacco usage among women has increased by 4.16% in the last 5 years.[37]

Further, we estimated CTU in SLT and smoke form to be around 20.89 and 18.91% respectively in India during the 2010 – 2022 periods. GATS 2 survey reports a similar 21.4% prevalence of SLT usage in Indian adults, whereas usage in smoke form (10.7%) was lesser than our estimate.[29] Furthermore, our estimate of SLT usage is in line with a meta-analysis conducted by Naznin et al. 2020 which was conducted using evidence from publications up until February 2019 and was exclusive to the use of SLT in Bangladesh and India. They utilized regional studies as well as studies and reports based on secondary data, and nationalized surveys. According to the study, the pooled prevalence of SLT usage was 25% and 22%, respectively.[38]

In this review, there is a wide disparity between our overall CTU estimate (35.25%) and individual SLT or smoke form usage estimate. Though our SLT/ smoke form usage estimate aligns well with recent nationalized survey reports, we acknowledge that the inadequacy of tobacco type-specific usage data among included studies may have underestimated our tobacco type-specific estimates. (i.e., only nine and ten studies respectively had SLT & smoke–tobacco type-specific usage estimates, further, some studies which otherwise reported higher overall CTU, did not provide type-specific data and thereby had to be excluded while estimating the pulled prevalence of tobacco type-specific usage.)

### Strength

In addition to estimating overall CTU among the adult general population in India, we presented prediction intervals that account for between-study heterogeneity. Even though the interval reported in this analysis is extremely broad (3.19 - 78.74), it gives light on the likelihood of another future study reporting a CTU nearing the upper confidence limit among similar regional populations.[39, 40]

GATS being a nationwide survey had some methodological concerns like under-sampling in high tobacco-consuming states based on Census 2011, representation of institutionalized population and migrants as GATS being a survey on household population, etc.[41] This review is a comprehensive evidence synthesis from diverse original studies across India.

The novelty of this report lies in including different study populations belonging to different administrative zonal locations and helps to achieve pooled estimates while accounting for those variations. The inclusion of pooled tobacco usage data from different zonal locations is beneficial to the policymakers as this provides a better picture of the tobacco usage burden at local sites which remains under-presented through existing national-level surveys.[10, 11]

### Limitations

Though we searched PubMed, Scopus, Science Direct, CINAHL and Google Scholar systematically and meticulously, we acknowledge that we may have missed a few potential studies that could be found in other digital databases.

The results of this study, however, should be interpreted with the limitations in mind, which include high heterogeneity due to differences in sampling, as well as the use of different definitions for current tobacco users across each included study. In terms of extrapolation of results in the broader population, our inclusion criteria were homogenous, though heterogeneity between studies exists in terms of individual study’s methodological differences, which may restrict some generalizability. We were unable to provide age and socioeconomic stratum-specific stratified CTU estimates due to the lack of extractable data in several included studies. We have not considered the quantity or duration of consumption, but instead presented estimates based on current/daily tobacco use.

Another major limitation of this review is that due to the unavailability of data we could not show representative estimates on CTU among transgender, intersex, and other non-binary identities, who were regarded as ‘Other gender’ than binary male/ female format in the last 2011 census in India. Keeping in mind, that around 30 lakhs (3 million) third gender population lives in India, future studies should explore the extent of tobacco usage among the population.

## Conclusion

Despite advances in Tobacco control efforts, its consumption continues to be a public health threat. In all administrative zones of India usage of Tobacco was prevalent, especially in East and North East regions coming at the top. Usage among women has also been seeing an increasing trend across the country with a slow or almost plateaued descent amongst men. These demand reinforcement in the form of institutional prevention programs, more aggressive risk communication, education, and most importantly, stringent national and regional policy enforcement. Further, this review highlights the need for more regionally representative studies alongside nationalized surveys to achieve more robust usage estimates.

## Data Availability

All data produced in the present study are available upon reasonable request to the authors

## Acknowledgment

We want to convey our gratitude to the previous peer reviewers of this manuscript whose insights have been crucial for technical improvement.

## Funding

No funding has been received for the conduction of this meta-analysis.

## Declarations of interest

None.

## Author Contributions

The authors confirm their contribution to the paper as follows: study conception and design: RT, SP. data collection: SP, DB, RT; analysis and interpretation of results: RT, SP; draft manuscript preparation: RT, SP. All authors reviewed the manuscript and have approved the final version of the manuscript.

## Data availability

Data used in this review is accessible and available in an open-source platform in the form of individual published studies. Compiled data can be shared upon request to the corresponding author through email communications.

## Notes

### Competing Interest Statement

The authors have declared no competing interest.

### Funding Statement

This study did not receive any funding

### Author Declarations

All the details of the studies included in the mnuscript has been mentioned in the manuscript

## References

[1] Danaei G, Vander Hoorn S, Lopez AD, et al. Causes of cancer in the world: comparative risk assessment of nine behavioural and environmental risk factors. Lancet 2005; 366: 1784–1793.

[2] Sirur S. 27.1% of India’s all cancer cases in 2020 will be tobacco-related, ICMR report estimates. ThePrint, https://theprint.in/india/27-1-of-indias-all-cancer-cases-in-2020-will-be-tobacco-related-icmr-report-estimates/484724/ (2020, accessed 29 December 2022).

[3] Mathers CD, Loncar D. Projections of global mortality and burden of disease from 2002 to 2030. PLoS Med 2006; 3: e442.

[4] GBD 2019 Tobacco Collaborators. Spatial, temporal, and demographic patterns in prevalence of smoking tobacco use and attributable disease burden in 204 countries and territories, 1990-2019: a systematic analysis from the Global Burden of Disease Study 2019. Lancet 2021; 397: 2337–2360.

[5] World Health Organization. WHO global report on trends in prevalence of tobacco use 2000-2025. 3rd ed. Geneva: World Health Organization, https://apps.who.int/iris/handle/10665/330221 (2019, accessed 29 December 2022).

[6] Sharapova S, Reyes-Guzman C, Singh T, et al. Age of tobacco use initiation and association with current use and nicotine dependence among US middle and high school students, 2014-2016. Tob Control 2020; 29: 49–54.

[7] NCD Alliance. Tobacco Use. NCD Alliance, https://ncdalliance.org/why-ncds/risk-factors-prevention/tobacco-use (2015, accessed 29 December 2022).

[8] Mercadal T. World Health Organization Framework Convention on Tobacco Control. In: The SAGE Encyclopedia of Cancer and Society. Thousand Oaks,: SAGE Publications, Inc., pp. 1325–1326.

[9] John RM, Sinha P, Munish VG, et al. Economic Costs of Diseases and Deaths Attributable to Tobacco Use in India, 2017–2018. Nicotine & Tobacco Research 2021; 23: 294–301.

[10] Shaikh R, Janssen F, Vogt T. The progression of the tobacco epidemic in India on the national and regional level, 1998-2016. BMC Public Health 2022; 22: 317.

[11] Rani M, Bonu S, Jha P, et al. Tobacco use in India: prevalence and predictors of smoking and chewing in a national cross sectional household survey. Tob Control 2003; 12: e4.

[12] van der Heijden TGW, Chilunga FP, Meeks KAC, et al. The Magnitude and Directions of the Associations between Early Life Factors and Metabolic Syndrome Differ across Geographical Locations among Migrant and Non-Migrant Ghanaians—The RODAM Study. IJERPH 2021; 18: 11996.

[13] Bethel AC, Rogers M, Abbott R. Use of a search summary table to improve systematic review search methods, results, and efficiency. J Med Libr Assoc; 109: 97–106.

[14] Munn Z, Stern C, Aromataris E, et al. What kind of systematic review should I conduct? A proposed typology and guidance for systematic reviewers in the medical and health sciences. BMC Med Res Methodol 2018; 18: 1–9.

[15] Palipudi KM, Morton J, Hsia J, et al. Methodology of the Global Adult Tobacco Survey — 2008– 2010. Glob Health Promot 2016; 23: 3–23.

[16] Study Quality Assessment Tools | NHLBI, NIH, https://www.nhlbi.nih.gov/health-topics/study-quality-assessment-tools (accessed 26 July 2022).

[17] RStudio Team (2022). RStudio: Integrated Development Environment for R. RStudio, http://www.rstudio.com/ (2022, accessed 30 September 2022).

[18] Barker TH, Migliavaca CB, Stein C, et al. Conducting proportional meta-analysis in different types of systematic reviews: a guide for synthesisers of evidence. BMC Med Res Methodol 2021; 21: 189.

[19] Nyaga VN, Arbyn M, Aerts M. Metaprop: a Stata command to perform meta-analysis of binomial data. Archives of Public Health 2014; 72: 39.

[20] Zeng W, Chen R, Wang X, et al. Prevalence of mental health problems among medical students in China. Medicine (Baltimore) 2019; 98: e15337.

[21] Higgins JPT, Thompson SG, Deeks JJ, et al. Measuring inconsistency in meta-analyses. BMJ 2003; 327: 557–560.

[22] Spineli LM, Pandis N. Prediction interval in random-effects meta-analysis. American Journal of Orthodontics and Dentofacial Orthopedics 2020; 157: 586–588.

[23] Hunter JP, Saratzis A, Sutton AJ, et al. In meta-analyses of proportion studies, funnel plots were found to be an inaccurate method of assessing publication bias. J Clin Epidemiol 2014; 67: 897–903.

[24] Lin L, Chu H. Quantifying Publication Bias in Meta-Analysis. Biometrics 2018; 74: 785–794.

[25] Willis BH, Riley RD. Measuring the statistical validity of summary meta-analysis and meta-regression results for use in clinical practice. Stat Med 2017; 36: 3283–3301.

[26] Sommer AE, Golden BP, Peterson J, et al. Hospitalized Patients’ Knowledge of Care: a Systematic Review. J Gen Intern Med 2018; 33: 2210–2229.

[27] WHO-Global Health Observatory Data. Prevalence of current tobacco use - South Asia, https://data.worldbank.org/indicator/SH.PRV.SMOK?locations=8S (2022, accessed 25 September 2022).

[28] Gururaja B R. Tobacco use down, chewing gutka on the rise in Karnataka, NFHS survey shows. Deccan Herald, May 2022, https://www.deccanherald.com/state/top-karnataka-stories/tobacco-use-down-chewing-gutka-on-the-rise-in-karnataka-nfhs-survey-shows-1113849.html (May 2022, accessed 25 September 2022).

[29] Tata Institute of Social Sciences (TISS), Mumbai and Ministry of Health and Family Welfare,, Government of India. Global Adult Tobacco Survey: India 2016-17 Report. TISS, Mumbai, https://www.tiss.edu/view/11/research-projects/global-adult-tobacco-survey-round-2-for-india-2016/.

[30] Rai B, Bramhankar M. Tobacco use among Indian states: Key findings from the latest demographic health survey 2019–2020. Tob Prev Cessat 2021; 7: 19.

[31] Suliankatchi Abdulkader R, Sinha DN, Jeyashree K, et al. Trends in tobacco consumption in India 1987–2016: impact of the World Health Organization Framework Convention on Tobacco Control. Int J Public Health 2019; 64: 841–851.

[32] Bapat, Gaikwad, Bramhankar, et al. Tobacco consumption declines across states, dry Bihar consumes more alcohol than Maharashtra: NFHS-5. Down-To-Earth, December 2020, https://www.downtoearth.org.in/blog/health/tobacco-consumption-declines-across-states-dry-bihar-consumes-more-alcohol-than-maharashtra-nfhs-5-74844 (December 2020, accessed 25 September 2022).

[33] International Institute for Population Sciences (IIPS) and ICF, 2021. National Family Health Survey (NFHS-5), 2019-21: Volume I, http://rchiips.org/nfhs/NFHS-5Reports/NFHS-5_INDIA_REPORT.pdf (2022).

[34] Sreeramareddy CT, Pradhan PMS, Mir IA, et al. Smoking and smokeless tobacco use in nine South and Southeast Asian countries: prevalence estimates and social determinants from Demographic and Health Surveys. Population Health Metrics 2014; 12: 22.

[35] Mackay J, Eriksen MP. The Tobacco atlas. Geneva: World Health Organization, 2002.

[36] Goel S, Tripathy JP, Singh RJ, et al. Smoking trends among women in India: Analysis of nationally representative surveys (1993–2009). South Asian J Cancer 2014; 3: 200–202.

[37] kadilal et al. A. Tobacco consumption on the rise in rural women: Report. Deccan Herald, https://www.deccanherald.com/state/tobacco-consumption-on-the-rise-in-rural-women-report-1037010.html (2021, accessed 29 September 2022).

[38] Naznin E, Wynne O, George J, et al. Systematic review and meta-analysis of the prevalence of smokeless tobacco consumption among adults in Bangladesh, India and Myanmar. Tropical Medicine & International Health 2020; 25: 774–789.

[39] Nagashima K, Noma H, Furukawa TA. Prediction intervals for random-effects meta-analysis: A confidence distribution approach. Stat Methods Med Res 2019; 28: 1689–1702.

[40] Faggion Jr CM, Menne MC, Pandis N. Prediction intervals should be included in meta-analyses published in dentistry. European Journal of Oral Sciences 2021; 129: e12827.

[41] Lal P, Srinath S, Goel S, et al. Unravelling India’s tobacco epidemic – priorities and recommendations for the second round of Global Adult Tobacco Survey (GATS). Glob Health Promot 2015; 22: 7–19.

