## Supplementary Material for "Tobacco usage among general Indian population: A meta-analysis of evidence drawn from regional studies between 2010 - 2022"

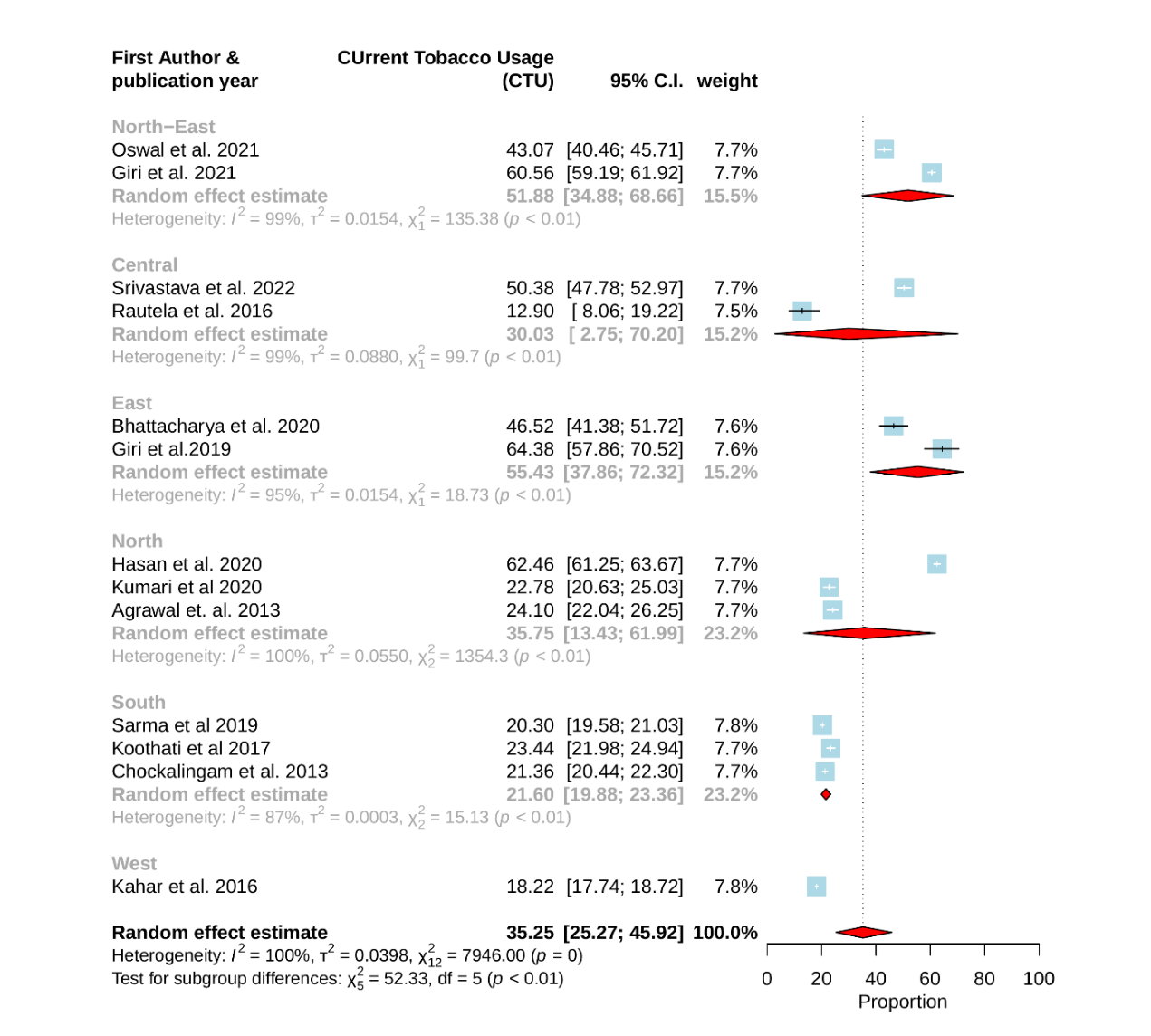
**Title: Tobacco usage among general Indian population: A meta-analysis of evidence drawn from regional studies between 2010 - 2022**

**Figure 1**: Forest plot of Subgroup analysis based on administrative Zone wise current tobacco usage among Indian adults (Number of studies included in analysis: 13)

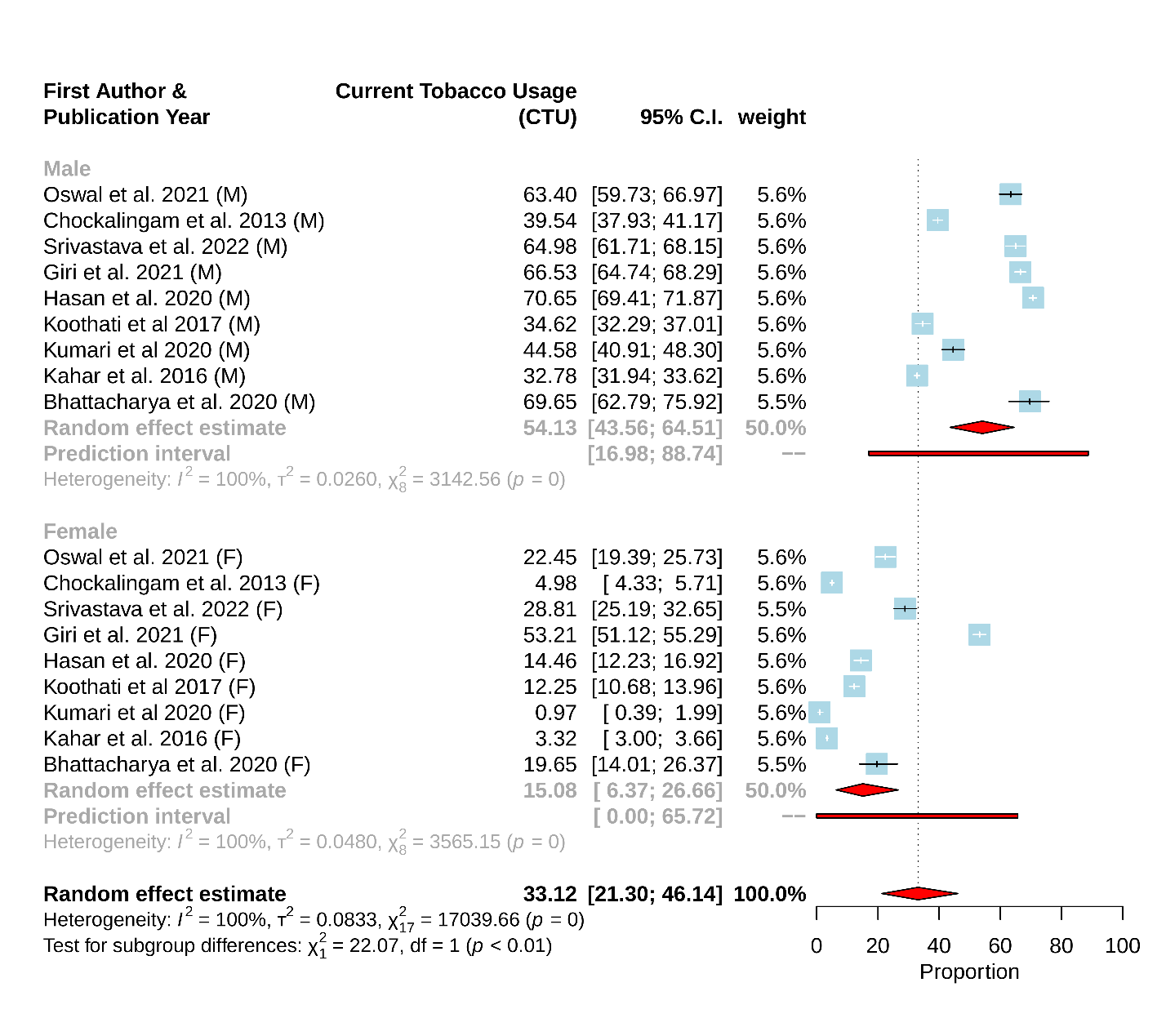
***Figure 2****: Forest plot of Subgroup analysis based on gender wise current tobacco usage among Indian adults (Number of studies included in analysis: 9)*

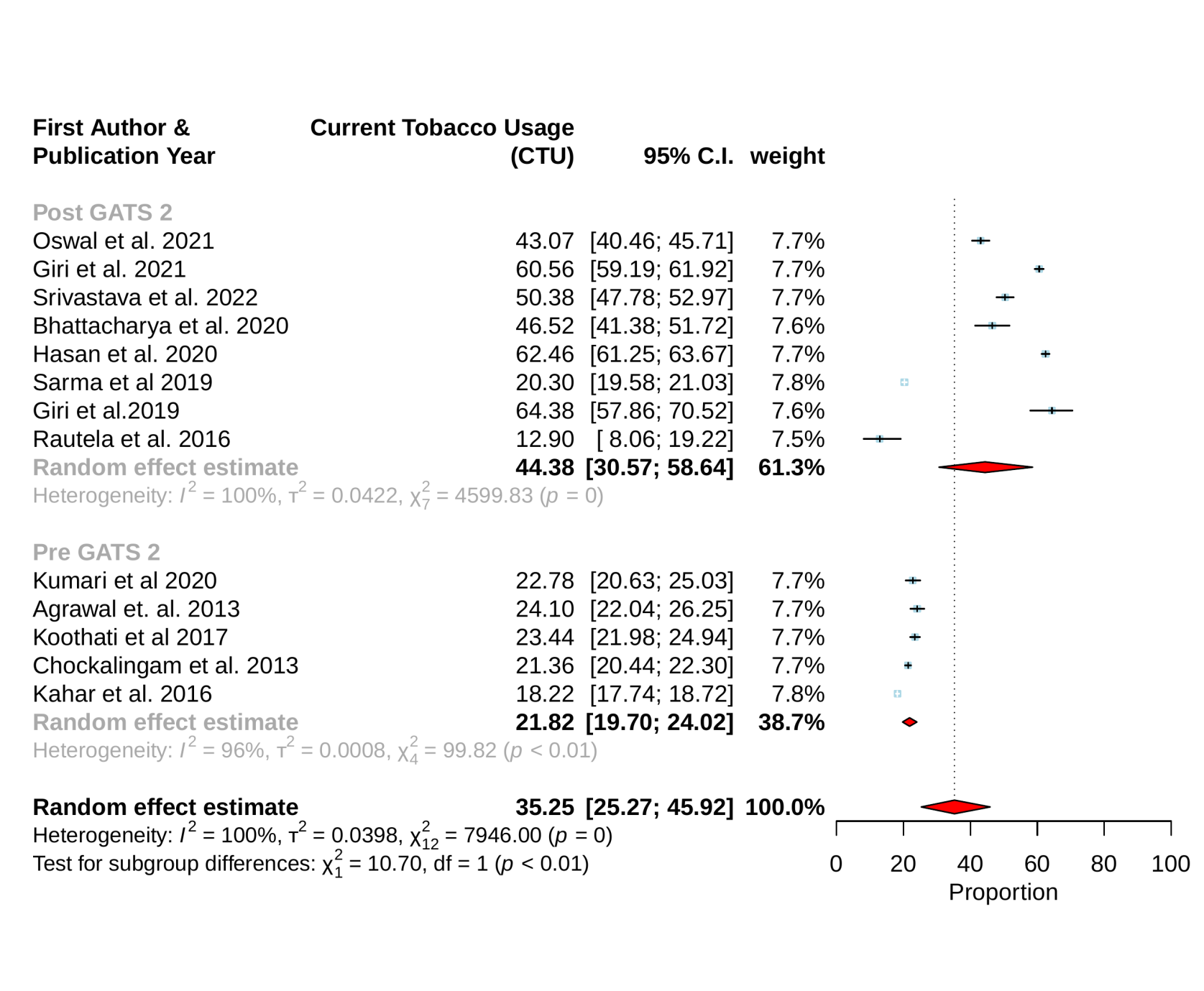

Figure 3: Forest plot of Subgroup analysis based on year wise current tobacco usage among Indian adults (Pre- GATS = 2010 – 2015, Post GATS includes GATS survey years till 2022) (Number of studies included in analysis: 13)
